# Enhancing multi-center generalization of machine learning-based depression diagnosis from resting-state fMRI

**DOI:** 10.1101/19004051

**Authors:** Takashi Nakano, Masahiro Takamura, Naho Ichikawa, Go Okada, Yasumasa Okamoto, Makiko Yamada, Tetsuya Suhara, Shigeto Yamawaki, Junichiro Yoshimoto

**Affiliations:** Division of Information Science, Graduate School of Science and Technology, Nara Institute of Science and Technology, Ikoma, 630-0192, Japan; Department of Psychiatry and Neurosciences, Hiroshima University, Hiroshima, 734-8551, Japan; Institute of Quantum Life Science, National Institutes for Quantum and Radiological Science and Technology, Chiba, 263-8555, Japan; Department of Functional Brain Imaging, National Institute of Radiological Sciences, Quantum Medical Science Directorate, National Institutes for Quantum and Radiological Science and Technology, Chiba, 263-8555, Japan

**Keywords:** depression, functional connectivity, machine learning, harmonization, multi-center fMRI, resting state fMRI

## Abstract

Resting-state fMRI has the potential to find abnormal behavior in brain activity and to diagnose patients with depression. However, resting-state fMRI has a bias depending on the scanner site, which makes it difficult to diagnose depression at a new site. In this paper, we propose methods to improve the performance of the diagnosis of major depressive disorder (MDD) at an independent site by reducing the site bias effects using regression. For this, we used a subgroup of healthy subjects of the independent site to regress out site bias. We further improved the classification performance of patients with depression by focusing on melancholic depressive disorder. Our proposed methods would be useful to apply depression classifiers to subjects at completely new sites.

## Introduction

Depressive disorder is a mental disorder characterized by long-lasting low mood. The diagnosis of depressive disorder has traditionally been made through the interaction between patients and doctors. It is important to develop more objective ways to diagnose depressive disorder in order to increase the reliability and accuracy of the diagnosis.

The combination of machine learning and functional magnetic resonance imaging (fMRI) has been used to diagnose or to find the physiological characteristics of psychiatric disorders [1-11].

Recently, functional connectivity (the correlation coefficients of the brain activity between brain regions) easily calculated from resting-state fMRI data are being used for the diagnosis of psychiatric disorders, such as autism, obsessive-compulsive disorder, and depression [1-7]. For resting-state fMRI, spontaneous brain activity was measured from subjects lying in an fMRI scanner without any stimulations. Because of these advantages of resting-state fMRI, functional connectivity has the potential to become a clinical tool in wide-spread hospitals.

However, functional connectivity is affected by site bias [2, 12]. To overcome site bias, using data from as many multiple sites as possible for the training of classification algorithms can be effective. Even though data from many sites were used for machine learning, it is difficult to apply this to a completely new site data.

In addition to site bias, the heterogeneity of major depressive disorder would be a problem when the classifier is applied to new data. Abnormality of functional connectivity of depression might be different depending on the subtype of the depression. It would be possible to improve classification performance by focusing on a typical severe depression called melancholic depressive disorder. In this paper, we propose the methods to improve the performance of diagnosis of major depressive disorder (MDD) at an independent site by reducing the site bias effects. In addition, we investigated the performance depending on the classification algorithms and the heterogeneity of the major depressive disorder.

## Materials and Methods

### Subjects

One hundred sixty-three patients with MDD (age 20-75, average 44.1 ± 12.2) were recruited by 5 sites (the Psychiatry Department of Hiroshima University and collaborating medical institutions, Table 1). They were screened using the Mini International Neuropsychiatric Interview (M.I.N.I,) [13, 14], which enables medical doctors to identify psychiatric disorders according to DSM-IV criteria [15]. Patients had an initial MRI scan before or after starting medication within 0-2 weeks.

**Table 1.** Demographic data of study participants.

| **Site 1 (HUH: Hiroshima University Hospital)** |  |  |  |
| --- | --- | --- | --- |
|  | HC | MDD | p value |
| No. of participants (Male/Female) | 59 (26/33) | 59 (32/27) |  |
| No. of melanchoria (Male/Female) | NA | 49 (29/20) |  |
| Age (years) | 33.7 ± 12.5 | 42.8 ± 12.0 | <0.001 |
| Severity of depression (BDI-II) | 6.9 ± 5.9 | 30.1 ± 8.5 | <0.001 |
| IQ (JART) | 113.7 ± 8.3 | 108.7 ± 9.7 | 0.004 |
| **Site 2 (HRC: Hiroshima Rehabilitation Center)** |  |  |  |
|  | HC | MDD | p value |
| No. of participants (Male/Female) | 12 (3/9) | 12 (6/6) |  |
| No. of melanchoria (Male/Female) | NA | 6 (0/6) |  |
| Age (years) | 42.4 ± 9.4 | 41.8 ± 10.4 | 0.667 |
| Severity of depression (BDI-II) | 11.0 ± 12.6 | 35.3 ± 10.0 | <0.001 |
| IQ (JART) | 111.5 ± 5.8 | 120.3 ± 5.1 | <0.001 |
| **Site 3 (HKH: Hiroshima Kajikawa Hospital)** |  |  |  |
|  | HC | MDD | p value |
| No. of participants (Male/Female) | 22 (5/17) | 22 (12/10) |  |
| No. of melanchoria (Male/Female) | NA | 14 (8/6) |  |
| Age (years) | 46.4 ± 9.4 | 44.0 ± 11.7 | 0.497 |
| Severity of depression (BDI-II) | 5.5 ± 5.3 | 29.8 ± 7.1 | <0.001 |
| IQ (JART) | 115.3 ± 5.7 | 117.8 ± 5.5 | 0.178 |
| **Site 4 (COI: Center of Innovation in Hiroshima University)** |  |  |  |
|  | HC | MDD | p value |
| No. of participants (Male/Female) | 55 (20/35) | 55 (24/31) |  |
| No. of melanchoria (Male/Female) | NA | 25 (11/14) |  |
| Age (years) | 53.7 ± 12.2 | 47.2 ± 13.0 | 0.011 |
| Severity of depression (BDI-II) | 8.5 ± 6.6 | 25.7 ± 9.6 | <0.001 |
| IQ (JART) | 107.4 ± 11.5 | 112.2 ± 10.4 | 0.020 |
| **Site 5 (QST: National Inst. for Quantum and Radiological Sci.and Tech.)** |  |  |  |
|  | HC | MDD | p value |
| No. of participants (Male/Female) | 47 (41/6) | 15 (9/6) |  |
| No. of melanchoria (Male/Female) | NA | 11 (6/5) |  |
| Age (years) | 24.4 ± 5.8 | 39.7 ± 10.3 | <0.001 |
| Severity of depression (BDI-II) | 4.6 ± 4.2 | 28.8 ± 10.2 | <0.001 |
| IQ (JART) | - | - | - |

HC, healthy control group; MDD, major depressive disorder patient group.
BDI-II, Beck Depression Inventory-II; JART, Japanese Adult Reading Test.
Values are presented as mean±s.d.

As a control group, 195 healthy subjects (age 20-75, average 39.1 ± 15.5) with no history of mental or neurological disease were recruited from the local community. All control subjects underwent the same self-assessment and examination administered to the MDD group. Thereafter, for the melancholic MDD classifier, the dataset was limited to have the subtype of melancholia (based on M.I.N.I.). The numbers of patients and healthy controls were set to be equal for each site except for the subjects from the National Institutes for Quantum and Radiological Science and Technology, in order to develop a classifier unbiased toward either group (see Table 1). The subjects from the National Institutes for Quantum and Radiological Science and Technology were used only for the test data (replication cohort).

The study protocol in this study was approved by the Ethics Committee of Hiroshima University and the Radiation Drug Safety Committee and by the institutional review board of the National Institutes for Quantum and Radiological Science and Technology, in accordance with the ethical standards laid down in the 1964 Declaration of Helsinki and its later amendments. All necessary patient/participant consent has been obtained and the appropriate institutional forms have been archived. The detailed properties of the subjects are shown in Table 1.

### Acquisition and preprocessing of functional MRI data

fMRI scanners were used to generate magnetic resonance images. Functional data were collected using gradient echo planar imaging (EPI) sequences. High-resolution T1-weighted magnetization-prepared rapid gradient echo images were also acquired before scanning the functional data. In the scan room with dimmed lights, participants were instructed not to think of anything in particular, not to sleep, and keep looking at a cross mark at the center of the monitor screen. The first 4 to 7 images were discarded to allow magnetization to reach equilibrium. All participants underwent an approximately 5 to 10 min resting-state scan. The scanners and imaging parameters are different depending on the site (see Table 1 and 2).

**Table 2.**
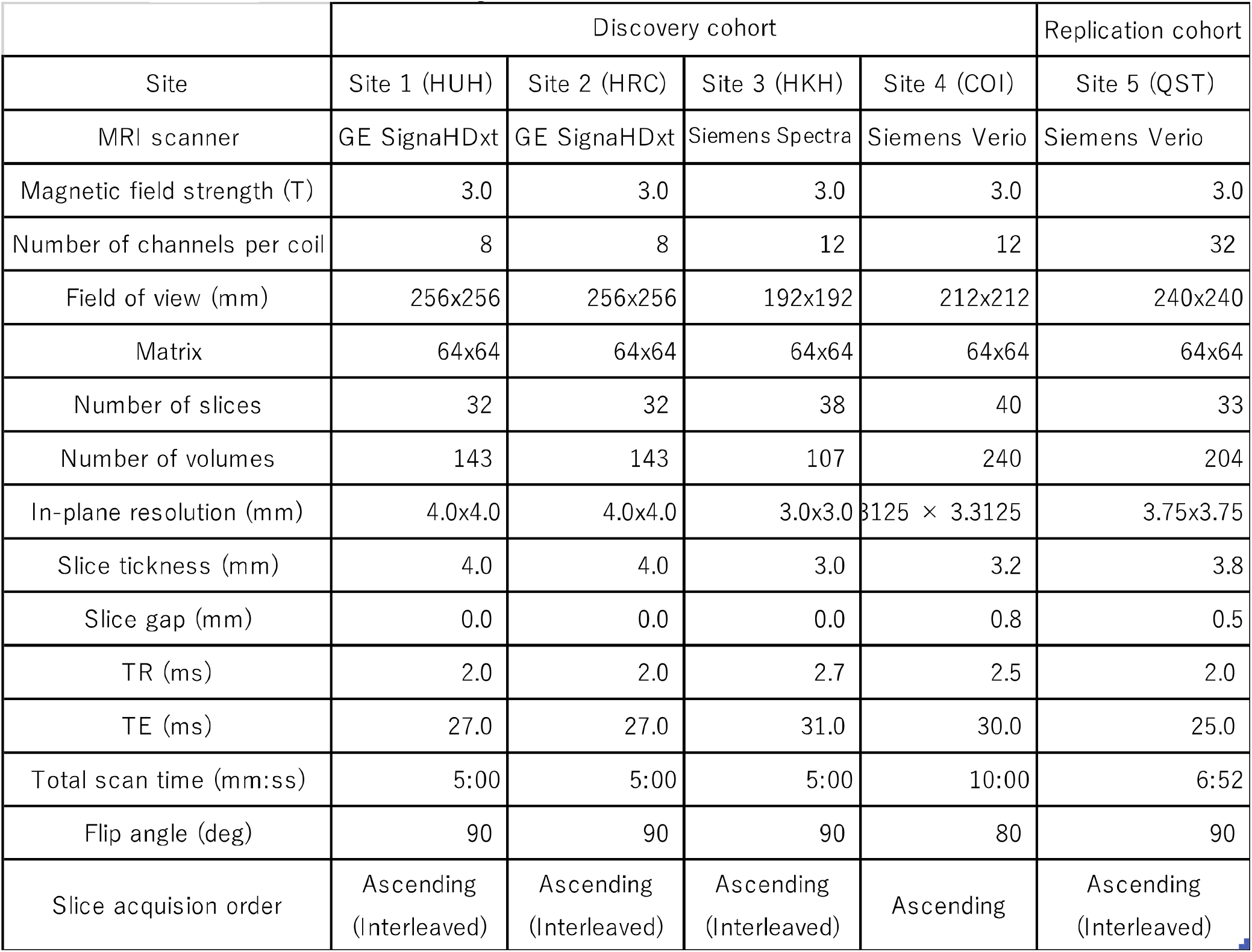
Imaging protocols for resting-state fMRI.

**Table 3.** Leave-One-Site-Out Cross-Validation

| site | site1 | site2 | site3 | site4 |
| --- | --- | --- | --- | --- |
| accuracy | 0.5 | 0.5 | 0.5 | 0.63 |
| sensitivity | 0 | 1 | 0 | 0.8 |
| specificity | 1 | 0 | 1 | 0.45 |

Subjects from three among four sites are used as the training data and their performance was evaluated using subjects from the remaining site. Area under the Receiver Operating Characteristic (ROC) Curve for Test Sites 1, 2, 3, and 4 are 0.71, 0.83, 0.66, and 0.73 respectively.

All the resting-state fMRI data were preprocessed using the identical procedures described in [7]. T1-weighted structural image and resting-state functional images were preprocessed using SPM8 (Wellcome Trust Centre for Neuroimaging, University College London, UK) on Matlab R2014a (Mathworks inc., USA). The functional images were preprocessed with slice-timing correction and realignment to the mean image. Thereafter, using the normalization parameters obtained through the segmentation of the structural image aligned with the mean functional image, the fMRI data was normalized and resampled in 2 ⨯ 2 ⨯ 2 mm^3^ voxels. Finally, the functional images were smoothed with an isotropic 6mm full-width half-maximum Gaussian kernel. After these preprocessing steps, the scrubbing procedure [16] was performed to exclude any volume (i.e., functional image) with excessive head motions, based on the frame-to-frame relative changes in time series data.

### Calculation of functional connectivity

For each individual, the time course of fMRI data was extracted for each of 137 regions of interest (ROIs), anatomically defined in the Brainvisa Sulci Atlas (BSA; http://brainvisa.Info) [6, 16] covering the entire cerebral cortex without a cerebellum. After applying a band-pass filter (0.008– 0.1 Hz), the following nine parameters were linearly regressed out: the six head motion parameters from realignment; the temporal fluctuation of the white matter; that of the cerebrospinal fluid; and that of the entire brain. Pair-wise Pearson correlations between 137 ROIs were calculated to obtain a matrix of 9,316 functional connectivities for each participant.

### Data standardization, removal of nuisance variables, and feature selections

For functional connectivity data, we applied the Fisher z-transformation to each correlation coefficient. We then regressed them on training subjects’ ages, sex, and dummy variables for each site. The resulting residuals of the regression are used for the subsequent procedures as the features controlling for age, sex, and site effects. We also used the regression coefficients to remove nuisance variables such as age, sex, and site from test subjects. Features like functional connectivities were high-dimensional. We then used a rank-sum test for dimensional reduction; we used the threshold of 0.05 unless noted. The procedures are summarized in Figure 1.

**Figure 1.**
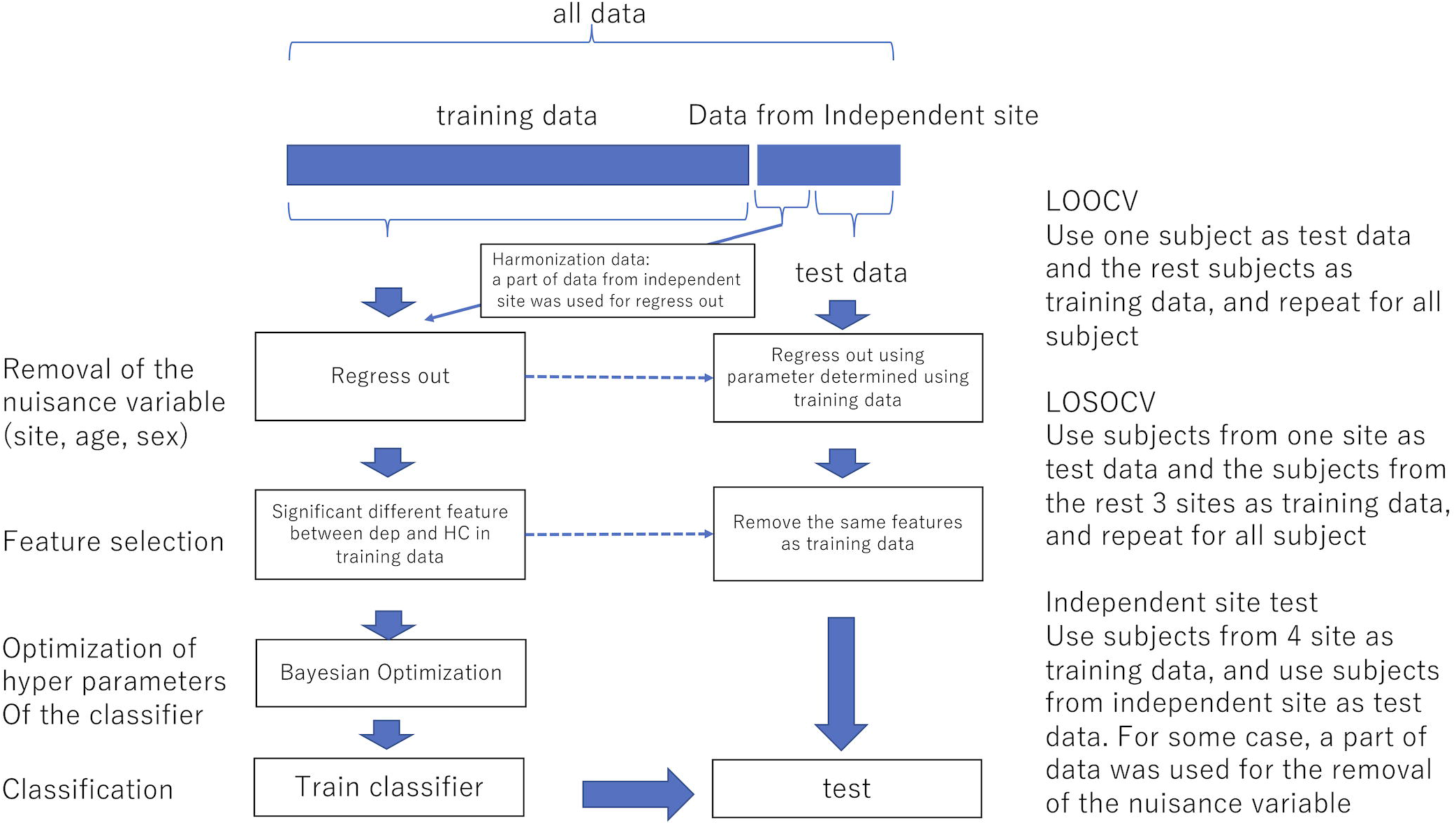
The procedure of the removal of site bias and feature selection. LOOCV Use one subject as test data and the remaining subjects as training data, and repeat for all subjects LOSOCV Use subjects from one site as test data and subjects from the remaining 3 sites as training data, and repeat for all subjects Independent site test Use subjects from four sites as training data, and use subjects from the independent site as test data. In some cases, a part of data was used for the removal of nuisance variables

### Classification algorithms

We then applied machine learning classification using the above features. We used ensemble learning (bagging and AdaBoost), support vector machine (SVM), or sparse logistic regression (SLR) [17] to classify depressed patients and healthy controls, or melancholic patients and healthy controls. Ensemble learning is a method to make a classifier by combining weak classifiers. While bagging makes the decision based on voting by parallel weak classifiers, AdaBoost combines weak classifiers sequentially. The hyperparameters adjusting tree depth (minimum leaf size and the maximum number of splits), and the other hyperparameters such as the number of ensemble learning cycles, and learning rates were optimized using Bayesian optimization described in the following section. Ensemble learning has advantages in dealing with small sample size, and high-dimensionality [18]. SVM is a widely used classifier and performs classification by finding the hyperplane that differentiates the two classes. Types of the kernel and penalty parameter were determined by the optimization in the following section. SLR is a Bayesian extension of logistic regression in which a sparseness prior is imposed on the logistic regression. SLR has the ability to select features objectively that are related to classifying depressive disorders [2, 7, 17].

### Evaluation and estimation of the hyperparameters of Classifiers

The performance of the classification was evaluated using Leave-One-Subject-Out cross-validation, Leave-One-Site-Out cross-validation within discovery cohort or independent dataset from a replication cohort.

For Leave-One-Subject-Out cross-validation, one subject was used for the test and the rest subjects were used for the training and this process was repeated until all subjects were used as a test data. The hyperparameters of the AdaBoost, Bagging, and SVM were optimized with Bayesian optimization using Gaussian process [19].

For the optimization, the whole data was divided into 9-folds keeping an equal amount of (diagnosis, gender, and site) combinations per fold. The optimization was performed using the 8-folds with 8-folds cross-validation (inner 7-folds are used for the training and the remaining inner one-fold was used for the validation). The test data was selected in the outer 1-fold. The rest from outer 1-fold and the outer 8-folds were used for the training of the classifier [7].

For Leave-One-Site-Out cross-validation, subjects from three out of the four sites were used as the training data and subjects from the remaining sites were used as test data. In the case that independent dataset was used as a test data, all subjects from the four sites were used for training. For both cases, the optimization of the hyperparameters was done using 8-folds cross-validation within the training data.

## Results

We trained classifier of the major depressive disorder using functional connectivity as the features. Because of the high dimensionality of functional connectivity, we selected the features using a rank-sum test with only the training data. Figure 2a shows the classification performance of AdaBoost, Bagging, SLR, and SVM with 0.05 threshold of the rank-sum test. while SVM shows the best classification accuracy, SLR did not show good accuracy. Then we investigated the classification performance such as accuracy, sensitivity (the number of subjects classified as patients divided by the number of patients) and specificity (the number of subjects classified as healthy controls divided by the number of healthy controls) and effect of threshold for the feature selection focusing on the SVM (Figure 2b). The classification performance of SVM was 73.3 % accuracy, 74.3 % sensitivity, and 72.3 % specificity with 0.05 threshold of the rank-sum test. The performance was not different even as we changed the threshold (Figure 2b).

**Figure 2.**
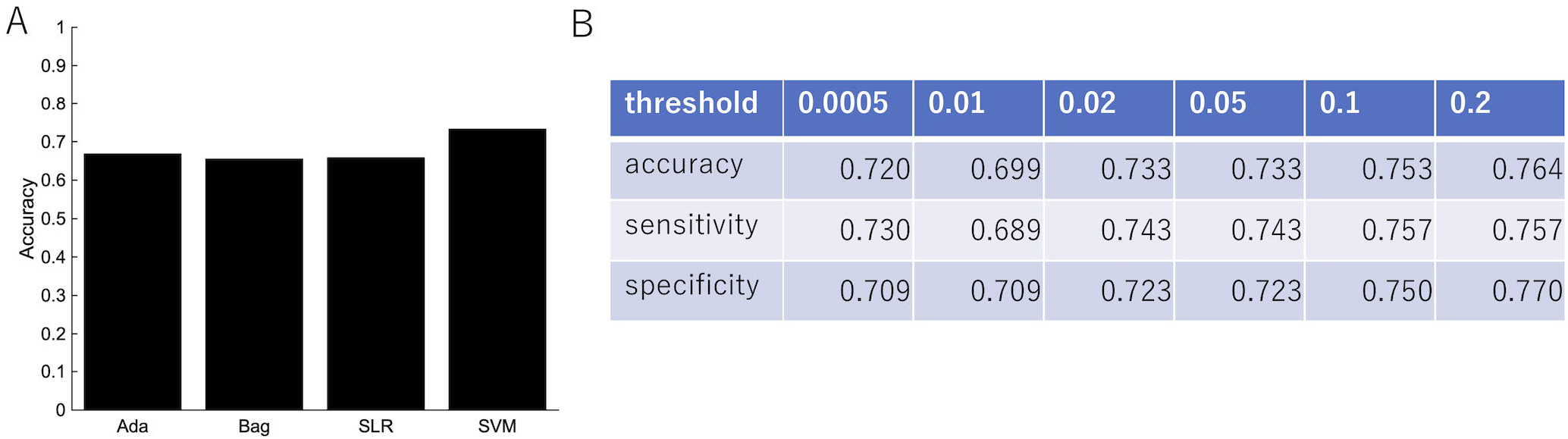
The performance of the classification of depressed patients. (a) The SVM shows an accuracy of 73.3%, sensitivity of 74.3%, and specificity of 72.3%. (b) The classification performance with different threshold of feature selection.

We then did a permutation test in order to confirm that the performance was significant. The diagnostic labels were randomly permuted and the performance of the classification was evaluated. The results showed that the classification accuracy was significantly higher than the change level for each algorithm (Figure 3).

**Figure 3.**
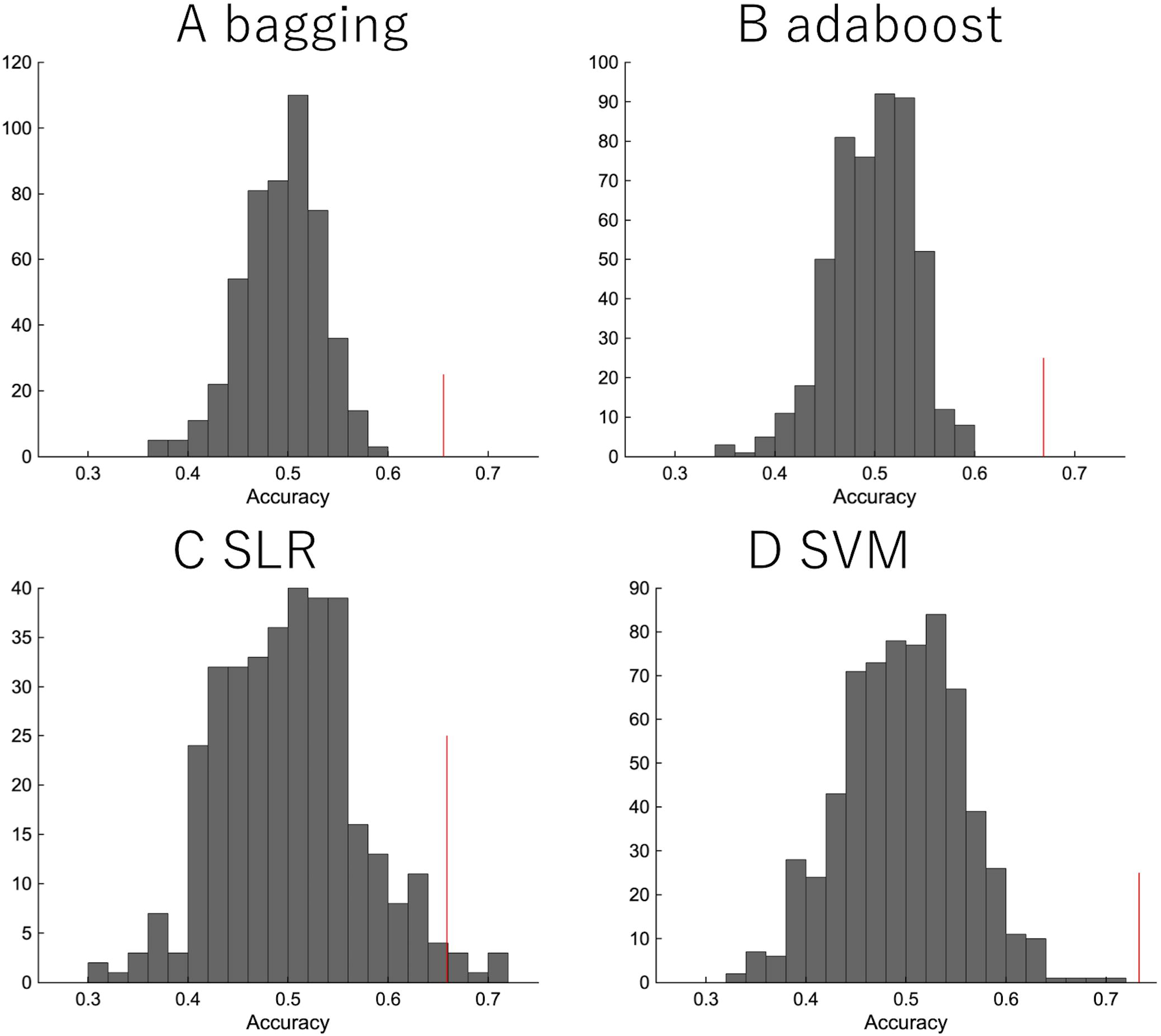
The permutation test. The diagnostic labels were randomly permuted for each subject and classification algorithms were applied, such as (a) random forest, (b) AdaBoost, (c) SLR, (d) SVM. The procedure was repeated 500 times.

In order to investigate the effects of site bias on the classification, the evaluation of classification was performed by leave-one-site-out cross-validation. The classification accuracies varied widely depending on the site (Table 2). Especially, the sensitivity and specificity were highly dependent on the site, which indicated that the classification was based on site bias rather than the feature associated with depression.

Depression is heterogeneous. This can be the reason why the classification performance was affected by the site rather than the diagnostic label. We then focused on melancholic depression that is a subtype of major depressive disorder with biological homogeneity [20-22]. The classification performance between melancholic patients and healthy controls was slightly higher than the one between major depressed patients, including non-melancholic patients and healthy controls (Figure 4). The classifier classified the patients with non-melancholic depression with an accuracy of about 75.3 %. The tendency of the accuracy depending on the algorithms was the same as in the case of the patients with major depressive disorder vs healthy controls. That is, SVM and ensemble learning showed good performance, whereas SLR did not. Moreover, the performance was evaluated by leave-one-site-out cross-validation. The classification accuracies varied widely depending on the site even without non-melancholic patients.

**Figure 4.**
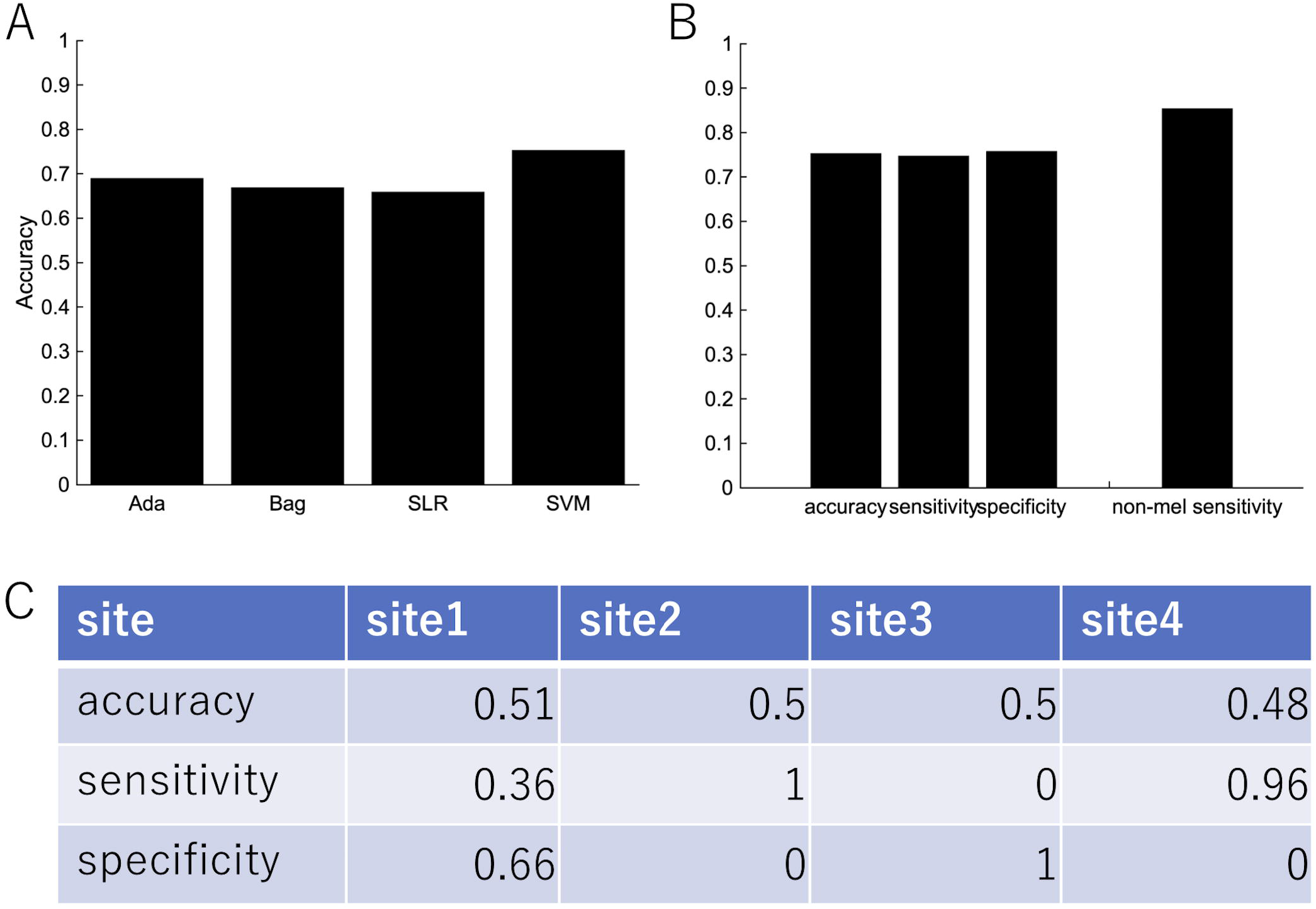
The performance of classification of melancholic patients and healthy controls. (a) Accuracy, sensitivity, and specificity of the random forest by LOOCV. The rightmost bar is the accuracy rate that the melancholic classifier correctly classifies non-melancholic patients as depressed patients. (b) LOOCV performance of melancholic patients’ classification using other classification algorithms. (c) Leave-one-site-out CV performance of melancholic patients’ classification. Area under the ROC Curve for Test Sites 1, 2, 3, and 4 are 0.66, 0.5, 0.82, and 0.79, respectively.

We then used subjects from the independent site (Figure 5). We trained the classifier using subjects from four sites and tested the classifier using subjects from the independent site. The performance shows the classification was based on site bias rather than the diagnostic label (Figure 5a). The decision boundary between patients and healthy controls obtained using training data is different from the boundary between patients and healthy controls of the test data. In order to overcome this problem, we used a part of the healthy controls from the independent site for regressing out the site, ages, and sex information, and the remaining healthy controls and depressed patients were used as test data. Note that the number of healthy controls and depressed patients used for the test data were set to be equal. Although the accuracy was 54.7%, this ingenuity reduced site bias (Figure 5b). Furthermore, when we focused on melancholic patients, the performance had drastically much improved (Figures 5d-f) and the accuracy was 71.9 %.

**Figure 5.**
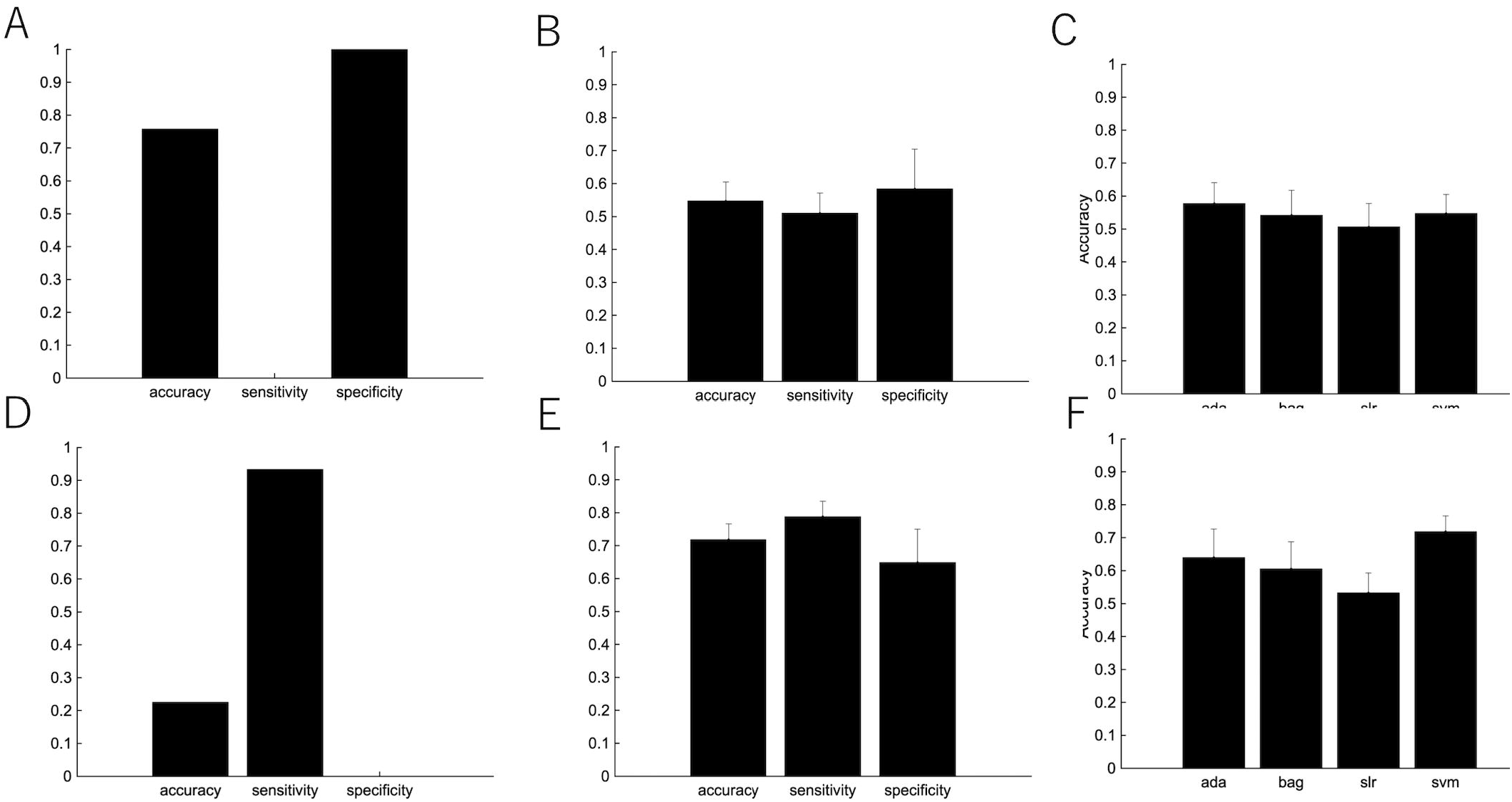
The improvement of the classification performance using the independent site. (a) All subjects from independent site data were used as test data. (b) The performance of the classification after part of healthy subjects (not used as either the training or test data) are used for the regressing out. Because there are many combinations from which to choose healthy controls used for the regressing out, we randomly chose healthy controls for the regressing out, which procedure was repeating 100 times. The error bar indicated the standard deviation. (c) Performance of several classification algorithms. (d-f) The same as above, but used only melancholic patients and healthy controls.

## Discussion

In this study, we proposed a method to reduce the site bias effect and improve the classification accuracy of patients with the major depressive disorder for an independent test dataset. The most effective improvement was achieved by the calibration using a part of the independent dataset. Specifically, a part of healthy controls was used for regressing out site bias. Furthermore, when we focused on the classification between melancholic depressed patients and healthy controls, the effect of the calibration was further improved. In this case, SVM showed the best performance.

### Generalization for different sites

There are some studies trying to overcome site bias. Some studies used independent component analysis or sparse canonical component analysis to obtain site-independent brain activity [23]. These methods are time-consuming and it is possible to lose important features that characterize patients with depression. On the other hand, regressing out is a simple way to reduce site bias and retains all features.

In our study, SVM showed good classification performance for independent site data. Depressive disorder is known to be heterogeneous. The melancholic depressive disorder is a typical and severe type of disorder. This would be a reason why the classification performance is better when we focus on melancholic patients. Targeting other types of depressive disorders would be future work. There are some studies about the subtyping the depressive disorder by using biological markers including functional connectivity [1, 6]. If a new subtype of depressive disorder that showed characteristic neural behavior is found, the diagnosis for this subtype using machine learning would be improved.

### For clinical diagnosis

When we consider an application for clinical diagnosis, site bias is a large problem. If traveling subjects, the same persons visit multiple sites, are available, the calibration would be more effective and better performance of the diagnosis of patients with major depression would be expected [2]. Even if traveling subjects are not available, our proposed procedure would be helpful. Site bias should be calibrated using data from healthy subjects before clinical diagnosis, using the fMRI scanner used for the clinical diagnosis, which would be a more practical way for an application at a novel site.

## Data Availability

The original data for this study is confidential due to the involvement of patient data. It can be obtained upon request to the Department of Psychiatry and Neurosciences, Hiroshima University, Japan (primary contact: Shigeto Yamawaki, PhD, MD). IRB imposing these restrictions on our data is Ethical Committee for Epidemiology of Hiroshima University (contact: Shoji Karatsu). Additional contact information is available from: https://www.hiroshima-u.ac.jp/en/pharm/contact.

## Acknowledgments

This research was supported by AMED under Grant Number JP19dm0107096, and Research on Medical ICT and Artificial Intelligence (H29-ICT-General-010), Health, Labour and Welfare Sciences Research Grants, Ministry of Health, Labour and Welfare Japan.

## Author Contributions Statement

T.N. and J.Y. carried out statistical analysis and gave interpretations. M.T., N.I., G.O., Y.O., M.Y., S.T., and S.Y. designed a study, collected functional MRI data. T.N and J.Y. wrote the main manuscript text and prepared the figures with input from all authors.

## Contribution to the Field Statement

An objective and accurate diagnosis method of depression is highly demanded to avoid misdiagnosis and suboptimal treatment outcomes. For the realization, resting-state functional magnetic resonance imaging (fMRI) combined with machine learning has recently attracted great attention. However, the potential measurement bias of fMRI derived from heterogeneous scanners and protocols makes it difficult to apply the classification method of depressive patients with depression at wide-spread hospitals. Therefore, even if a good classifier of the patients is created within a specific facility, the performance is deteriorated at other facilities. Here, we propose a method to overcome the problem by correcting the bias in advance using healthy persons in the new facility. In addition, by focusing only on the melancholy type, the classification performance at new facilities was improved. Our method is easy to implement at any facility, contributing to the diagnosis of depression at wide-spread hospitals.

